# What Specimen Urologists Should Be Most Concerned About ? A Systematic Review and Meta-Analysis

**DOI:** 10.1101/2020.10.08.20209544

**Authors:** M. Reza Roshandel, Masoud Nateqi, Ramin Lak, Pooya Aavani, Reza Sari Motlagh, Tannaz Aghaei Badr, John Sfakianos, Steven A. Kaplan, Shahrokh F. Shariat, Ashutosh K. Tewari

## Abstract

**Objective:** Investigating the infectivity of body fluid can be useful for preventative measures in the community and ensuring safety in the operating rooms and on the laboratory practices.

**Methods:** We performed a literature search of clinical trials, cohorts, and case series using PubMed/MEDLINE, Google Scholar, and Cochrane library, and downloadable database of CDC. We excluded case reports and searched all-language articles for review and repeated until the final drafting. The search protocol was registered in the PROSPERO database.

**Results:** Thirty studies with urinary sampling for viral shedding were included. A total number of 1,271 patients were enrolled initially, among which 569 patients had undergone urinary testing. Nine studies observed urinary viral shedding in urine from 41 patients. The total incidence of urinary SARS-CoV-2 shedding was 8%, compared to 21.3% and 39.5 % for blood and stool, respectively. The summarized risk ratio (RR) estimates for urine positive rates compared to the pharyngeal rate was 0.08. The pertaining RR urine compared to blood and stool positive rates were 0.20 and 0.33 respectively.

**Conclusions:** Our review concludes that not only the SARS-CoV-2 can be excreted in the urine in eight percent of patients but also its incidence may have associations with the severity of the systemic disease, ICU admission, and fatality rates. Moreover, the findings in our review suggest that a larger population size may reveal more positive urinary cases possibly by minimizing biases. However, it is important to notice that it is the naso-pharyngeal specimens, stool, and serum that show more possibilities to became positive, respectively.

**Take-home bullet points:**

❖ The **urinary shedding incidence was 8%,** compared to 21.3% and 39.5 % for blood and stool, respectively.
❖ Urinary shedding may have **associations with the severity** of the systemic disease, **ICU** admission, and **fatality** rates.
❖ **Repeat urinary testing is warranted throughout the disease phases**, especially in clinically suspected cases with an initially negative results.
❖ Technical errors in handling samples, as well as different rRT-PCR methods can be responsible for diversity found in results, in part.

## Introduction

Urinary viral shedding can be important from the aspects of diagnosis, vertical and horizontal transmission of infection (1). Viral shedding has been proven for some other contagious viruses including the *Ebola virus, Zika virus, and hepatitis-B virus* (2, 3).

Up to date, SARS-CoV-2 has spread to 213 countries and territories and infected over 28,000,000 patients around the globe with around 1,000,000 death toll (4). Before Coronavirus disease-2019 (COVID-19), the latest coronavirus outbreaks were the severe acute respiratory syndrome coronavirus-1 (SARS-CoV-1) and Middle Eastern respiratory syndrome coronavirus (MERS-CoV) outbreaks. For SARS-COV-1, the urinary positivity rates were reported to be up to 42% (5). SARS-CoV-2 structural features are similar to both SARS-CoV-1 and MERS-CoV which all belong to the family *Coronaviridae* (6).

The presence of SARS-CoV-2 has been shown in urine (7-12). Angiotensin-converting enzyme-II (ACE2) has been known as the cellular entry receptor of SARS‐ CoV‐2 (13, 14). The cells with ACE-II receptors such as epithelia of lung, kidney, and bladder may act as targets to SARS-CoV-2 (9, 15, 16). Although there are discrepancies in the reported results of the studies over the SARS CoV-2 urinary shedding, since the viral dynamics are yet to be fully determined, it’s been recommended that the urethral or ureteral instrumentation and handling should be carried out cautiously (17).

Determining whether the virus is detectable throughout the disease is critical to control transmission. Considering the stability of SARS-CoV-2 for up to 72 hours (18), performing urological surgeries, or collecting infected urinary samples may put urologists and health care workers at risk (19). The Ebola virus epidemic (2014 to 2015) was an awakening alarm for the health care community regarding the lack of biosafety in the handling of samples containing suspected special pathogens (20). This is true, particularly when responding to a not well-known pathogen, as the recommendations are often fluid (21). Learning about the infectivity also can alter preventative measures in the operating room and the settings needed for safety on laboratory practices (22).

Furthermore, the probability of transmission by pets (23) (24), leaves urine with a large potential to be a source of disease spread (25, 26). Although no data are available to confirm or exclude the possibility of such transmission, CDC advises restricting contact with pets and other animals while one has COVID-19 (27).

By this review, we systematically investigated the findings on the urinary SARS-Cov-2 to points out the important methodological considerations needed to be considered in future studies.

## Methods and materials

The protocol for this systematic review was registered with the International Prospective Register of Systematic Reviews (CRD42020187294). The review follows the PRISMA (Preferred Reporting Items for Systematic Reviews and Meta-analysis) statement (28).

### Literature Search Strategies

A systematic search of the literature was performed in PubMed/MEDLINE, Google Scholar, Cochrane library, and COVID-19 research articles downloadable database of CDC (Centers for Disease Control and Prevention). The comprehensive literature was performed in June 2020. No language restrictions were applied. Articles published in 2019 and 2020 were included. Searches were repeated until the final drafting of the manuscript, to capture emerging evidence from the ongoing studies. The searches included medical subject headings (MeSH) and keywords for SARS-CoV-2, COVID, Corona, together with shedding, persistence, urine, urinary, specimen, viral load, or RNA body fluids. Searches were designed to be broad and comprehensive initially, using the following keywords and MeSH terms: (“specimen” or “urine” or “urinary”) and (“corona” or “coronavirus” or “COVID” or “COVID-19” or “COVID-2019” or “SARS” or “SARS-CoV” or “SARS-CoV-2”).

### Eligibility criteria and study selection

Study selection was based on predefined eligibility criteria within a CoCoPop (Condition, Context, Population) and a PIRD (Population, Index Test, Reference test, Diagnosis of Interest) format (Table 1) (29). Additional exclusion criteria were applied at the full-text stage. After finalizing the selection of studies on the urinary SARS-CoV-2, information on viral existence in stool and blood specimens were explored in the selected studies (Figure 1).

**Table 1:** List of inclusion/exclusion criteria

|  | **Inclusion** | **Exclusion** |
| --- | --- | --- |
| **Population** | Any age with a confirmed COVID-19 on the admission or later | • Animal studies |
| **Interventions** | Real-time RT-PCR | • Limited to non-RT-PCR<br>• Innovative methods or uncommon genes incorporated to the test |
| **Comparisons** | Stool and serum specimens |  |
| **Outcomes** | Existence of shedding of viral RNA |  |
| **Type of Study** | Any study more equal/more than 2 cases with original data (including editorials, letters, comments, abstracts, summaries, Case reports ) | • Case Reports<br>• Review articles<br>• Publications with no original data (e.g. comments, reviews)<br>• Non-English publications |
| **Timing and Setting** | • 2019 and 2020<br>• All settings |  |

**Figure 1:**
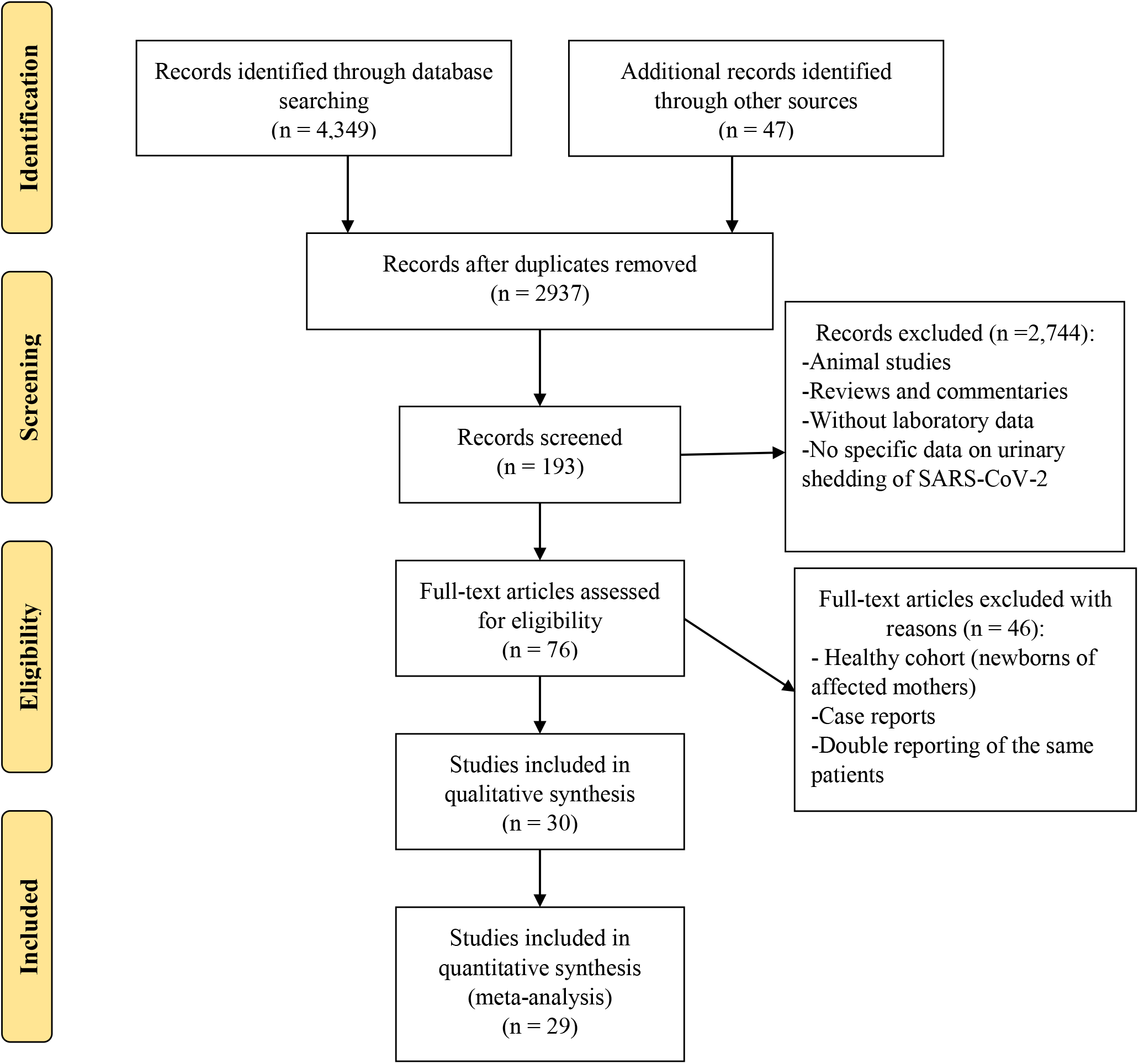
Preferred reporting items for systematic reviews and meta-analyses (PRISMA) flow diagram shows the study selection process. Adapted from Moher *et al*. (doi.org/10.1371/journal.pmed.1000097) ©2009, under terms of Creative Commons Attribution 4.0 International License.

All studies required a minimum of 2 patients were included. All potential studies were independently screened by two investigators. In the case disagreement, it resolved either through discussion or the involvement of a third researcher according to Delphi consensus criteria. Clinical trials, retrospective, prospective observational, case series, and cross-sectional studies were included as well as supplementary or non-peer-reviewed reports, correspondence, research letters, and preprints. Review articles, case reports, or non-relevant articles were excluded from the pool. Following reviewing and extraction of data, references of each manuscript were searched for relevant missing manuscripts.

### Data Extraction

We created standardized forms for data extraction and the pilot tested the forms before the process of data extraction. We completed the data abstraction process using created forms to record study characteristics, clinical data, and laboratory data including study year and design, country of study origin, total initial population size, test type for disease diagnosis, test type for samples (urine/stool/rectal swab/blood), patients age (including mean and range), number of positive and total patients and/or (wherever applicable) number of positive and total specimens collected for each test category, disease severity, ICU admission, and fatality rate. More details for study items are shown in Table 2.

**Table 2:** Comparison among studies with and without positive results on the urinary viral rRT-PCR

|  | Studies with positive findings | Studies with completely negative urine |
| --- | --- | --- |
| **Number of studies** | 9 studies | 21 studies |
| **Countries: number of studies** | 7: China<br>2: Korea | 11: China<br>2: USA<br>2: Germany<br>2: France<br>1: Iran<br>1: Singapore<br>2: Korea |
| **Initial number of enrolled patients** | 715 | 556 |
| **Final number enrolled for urinary testing** | 372 | 197 |
| **Stool positivity** | 42% | 42% |
| **Blood positivity** | 22% | 21% |
| **Severity of the disease** | 50.6% | 21% |
| **ICU** | 18% | 28.3% |

### Risk of Bias Assessment and Strength of Body of Evidence

Two investigators assessed the risk of bias for individual studies independently using JBI’s (Joanna Briggs) critical appraisal tools for prevalence study and diagnostic test accuracy studies to assess the trustworthiness, and results of the studies (30-32).

### Statistical Analysis

Forest plots were used to assess risk ratio (RR) and summarized them to describe RR of viral shedding rates in the urine and control groups (i.e., nasopharynx, stool, blood). Primary, secondary, and tertiary meta-analysis was conducted among all studies that reported urine and nasopharynx positive rates, urine, and stool positive rates, urine, and serum positive rates as an outcome, respectively. The heterogeneity across studies was evaluated using p-values, and Q and I2 statistics. Random effect and fixed effect meta-analysis were used when the heterogeneity was greater and lower than 50%,respectively. P-values lower than 0.05 were considered statistically significant. All analyses were carried out using Stata version 14.

## Results

A total of 30 studies met the inclusion criteria and were included in the review (8-13, 17, 35-49). The overall prevalence of urinary SARS-CoV-2 shedding was 8%. This was 21.3% and 39.5 % for blood and stool respectively.

### Study Characteristics and Urinary Testing Population

Characteristics of the included studies, comparison among positive and negative studies are detailed in Tables 2, 3, and Figure 2.

**Table 3:** Demographic information of studies and urinary viral results

| Study | Country | Initial population | Age (mean/median) | Patients<sup>¶</sup> with (+) urine among tested ones | Urine samples<sup>¶</sup> tested |
| --- | --- | --- | --- | --- | --- |
| Zhang N (33) | China/Beijing | 23 | 48 * | 2/23 | 2/56 |
| Ling Y (34) | China/Shanghai | 66 | 44 | 4/58 |  |
| Wang L (35) | China/Wuhan | 116 | 54 | 5/53 |  |
| Zheng S (10) | China | 96 | 55 | 1/67 |  |
| Tan W (36) | China | 67 | 49 | 12/64 |  |
| Mondanizadeh M (37) | Iran | 50 | 46 | 0/50 |  |
| Peng L (38) | China | 9 | 38.9 | 1/9 |  |
| Kujawski SA (39) | USA/CDC | 12 | 53 | 0/10 |  |
| Ghinai I (40) | USA/Illinois | 2 | NA | 0/2 | 0/12 |
| Young BE (41) | Singapore | 18 | 47 | 0/10 |  |
| Wölfel R (42) | Germany/Munich | 9 | NA | 0/9 | 0/27 |
| Lescure XF (43) | France/Paris | 5 | 48 | 0/5 |  |
| Yu F (44) | China/Beijing | 76 | 40* | NA | 0/14 |
| To KKW (45) | China/Hong Kong | 23 | 62* | 0/23 |  |
| Chan JFW (16) | China/Hong Kong | 5 | 55.4 | 0/5 |  |
| Wang W (46) | China/Beijing | 205 | 44 | NA | 0/72 |
| Pan Y(X) (47) | China/Beijing | 2 | NA | 0/2 |  |
| Lo IL (48) | China/Macau SAR | 10 | 54 | 0/10 | 0/49 |
| Chan JFW (49) | China/Hong Kong | 15 | 63 | NA | 0/33 |
| Chen Y (50) | China/Wuhan | 42 | 51 | 0/10 |  |
| Fang Z (51) | China/Xiangtan | 32 | 41 | 0/23 |  |
| Mumm JN (52) | Germany/Munich | 7 | 62 | 0/6 |  |
| Diao B (53) | China/Wuhan | 259 | NA | 14/19 |  |
| Qiu L (54) | China/Beijing | 10 | 52 to 80 | 0/10 |  |
| Xie C (55) | China/Chengdu | 19 | 33 | 0/9 |  |
| Kim JY (56) | Korea/ Seoul | 2 | 45 | 0/2 |  |
| Couturier A (57) | France/Paris | 2 | 53 | 0/2 |  |
| Jeong HW (58) | Korea/Cheongju | 5 | 63 | 5/5 |  |
| Liu P (59) | China/Shanghai | 9 | NA | 0/9 |  |
| Kim JM (60) | Korea/Cheongju | 74 | 43 | 2/74 | 2/274 |

\* median;
¶ the provided numbers are positive cases/specimens out of available total ones

**Figure 2:**
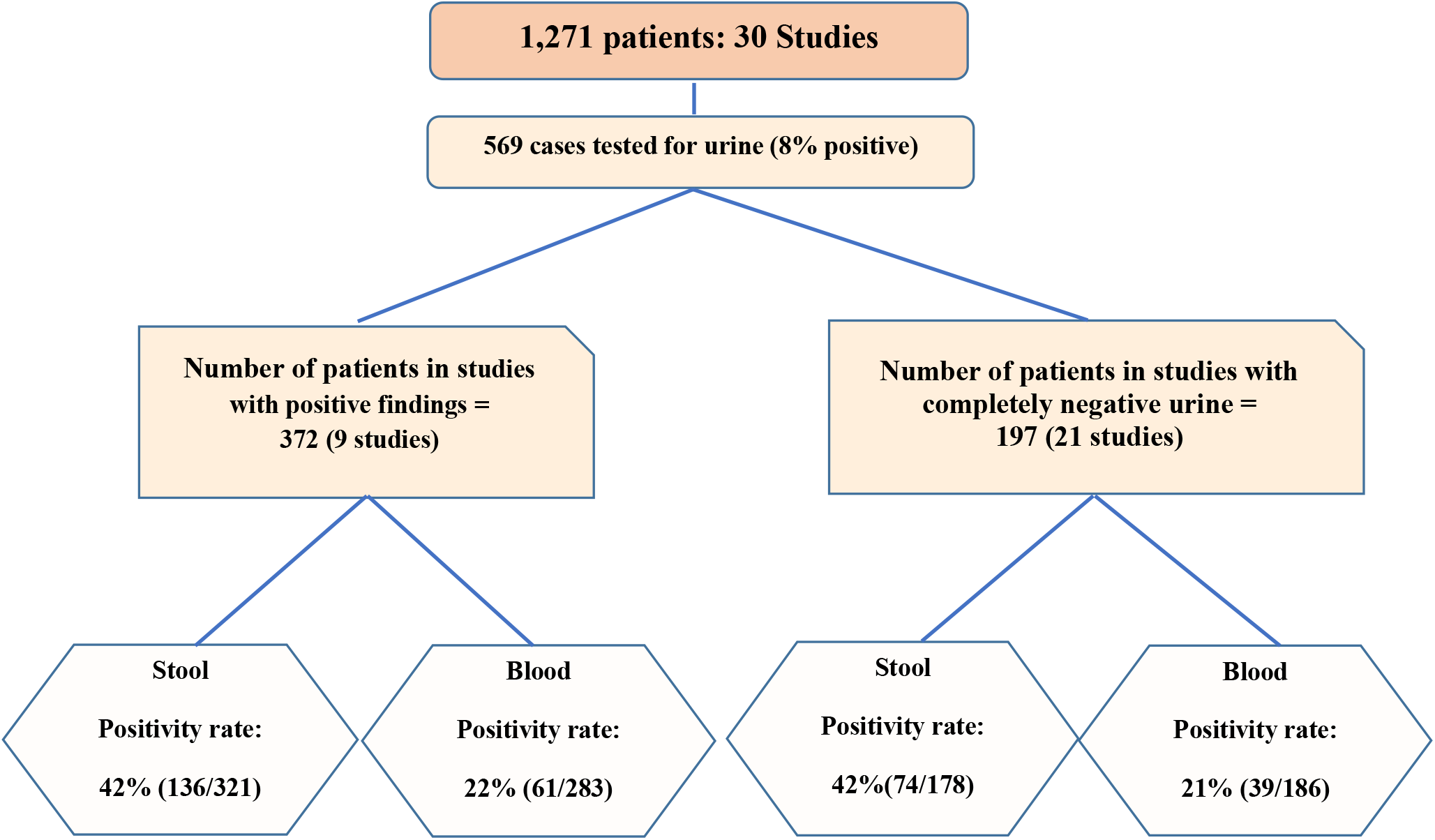
Flow chart diagram comparing studies in terms of urine, serum, and stool specimens

I. **Primary Study Population:** A total number of 1,271 COVID-19 patients were enrolled initially in the studies. The confirmatory testing was positive pharyngeal swabs, except two studies that included four patients with initially negative pharyngeal but positive stool results (13, 40). Essentially, all studies included patients with a median age between 40s and 60s (ranged from 6 months to 92 years).
II. **Ultimate Urinary Tested Population:** Of 746 patients, 439 had undergone urinary testing. In three studies, the number of urinary tests patients were not available, instead, the total number of laboratory samples (119 samples in total) was reported (Diagram 2).
III. **Stool/rectal Swab and Blood Sampling:** Of 30 studies with urinary sampling for viral shedding, 21 studies performed fecal/rectal swab testing and 22 studies accomplished blood viral testing.

### Laboratory Methods to Identify SARS CoV-19 in the Literature

Within the literature examined, the most commonly used assays for detection of SARS-CoV-2 in different samples such as urine, stool, blood, and pharyngeal swabs were RNA extraction followed by semi-quantitative and quantitative real-time reverse transcriptase-polymerase chain reaction (real-time RT-PCR, or rRT-PCR). In some studies, serological and molecular methods such as enzyme-linked immunosorbent assay (ELISA), partial and whole-genome sequencing were used for more verification. Several specific primers pairs were used to amplification of gene regions including RdRp/helicase, spike, and nucleocapsid genes of SARS-CoV-2. Such data were not available for all papers.

### Severity, ICU admission, Fatality rate, and patient enrolling

The COVID-19 severity and ICU admission rates are detailed in Table 4.

**Table 4:**
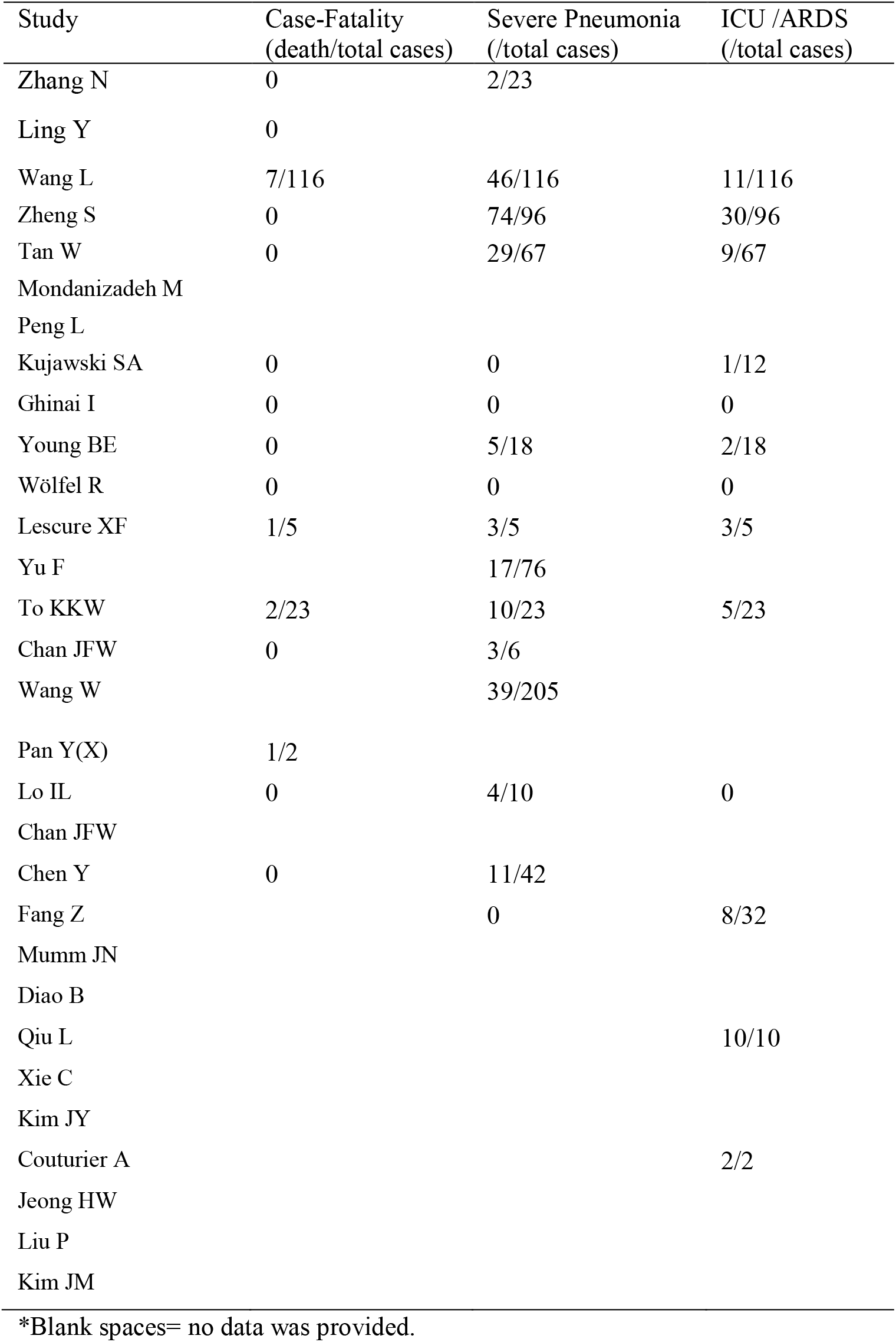
Clinical characteristics of patients in studies

- 20.1% of the total initial population were admitted into the ICU, as reported in 13 studies.
- 33.0% of the total initial population had severe respiratory disease because of SARS-CoV-2, as reported in 16 studies.
- There was no information about the severity or ICU admission in the rest of the studies. Four studies that included the cases terminated by death reported a total fatality rate of 7.6% (11/144). The other 11 studies only enrolled survived cases. Related information was not provided in 15 studies.

### Shedding in urine and other specimens

The results are shown in Table 5 and Figure 2.

**Table 5:** Positive real-time RT-PCR (rRT-PCR) findings of other specimens in the studies

| STUDY | Stool<sup>(+)</sup><br>Patients<sup>¶</sup> | Stool<br>(+)Specimens<sup>¶</sup> | Blood<sup>(+)</sup><br>Patients<sup>¶</sup> | Blood<sup>(+)</sup><br>Specimens<sup>¶</sup> |
| --- | --- | --- | --- | --- |
| Zhang N | 10/12 | 33/51 | 0/23 | 0/56 |
| Ling Y | 11/66 | - | 0/14 | - |
| Wang L | - | - | - | - |
| Zheng S | 55/93 | - | 39/95 | - |
| Tan W | 45/62 | - | 9/63 | - |
| Mondanizadeh M | 2/50 | - | 3/50 | - |
| Peng L | 2/9 | - | 2/9 | - |
| Kujawski SA | 7/10 | - | 1/12 | - |
| Ghinai I | 1/2 | 4/11 | 0/2 | 0/14 |
| Young BE | 4/8 | - | 1/12 | - |
| Wölfel R | 0/4 | 0/13 | 0/9 | 0/31 |
| Lescure XF | 2/5 | - | 1/5 | - |
| Yu F | - | - | NA | 0/4 |
| To KKW | 4/23 | - | 5/23 | - |
| Chan JFW | 0/2 | - | 2/6 | - |
| Wang W | NA | 44/153 | NA | 3/307 |
| Pan Y(X) | 0/2 | - | - | - |
| Lo IL | 10/10 | 46/79 | - | - |
| Chan JFW | NA | 7/33 | NA | 10/87 |
| Chen Y | 28/42 | - | - | - |
| Fang Z | - | - | 23/32 | - |
| Mumm JN | - | - | 2/4 | - |
| Diao B | - | - | - | - |
| Qiu L | - | - | 0/10 | - |
| Xie C | 8/9 | - | 0/9 | - |
| Kim JY | 0/2 | - | 1/2 | - |
| Couturier A | - | - | 0/1 | - |
| Jeong HW | 5/5 | - | 5/5 | - |
| Liu P | 8/9 | - | 0/9 | - |
| Kim JM | 8/74 | 13/129 | 6/74 | 9/323 |

¶ the provided numbers are positive cases/specimens among tested cases
\*rRT-PCR=Real-time Reverse Transcription Polymerase Chain reaction

I. Urinary results: Of 30 studies, urinary viral shedding was observed in nine studies.
II. Stool or rectal swab results: 19 of 23 studies found positivity in the stool samples.
III. Blood testing results: 14 out of 25 studies reported positive results.

### Meta-analysis

Meta-analysis was performed across the urinary studies. The summarized RR of 30 retrospective studies that reported urine compared to nasopharynx positive rates, was 0.08 (95% confidence intervals (CI); 0.05-0.16). The pertaining RR urine compared to blood and stool positive rates were 0.20 (95% CI; 0.14-0.29) and 0.33 (95% CI; 0.15-0.72) respectively. The forest plots of the meta-analysis are shown in Figures 3-5. There was no significant heterogeneity across all studies that included in the all meta-analysis, therefore fixed-effect analyses have been used.

**Figure 3:**
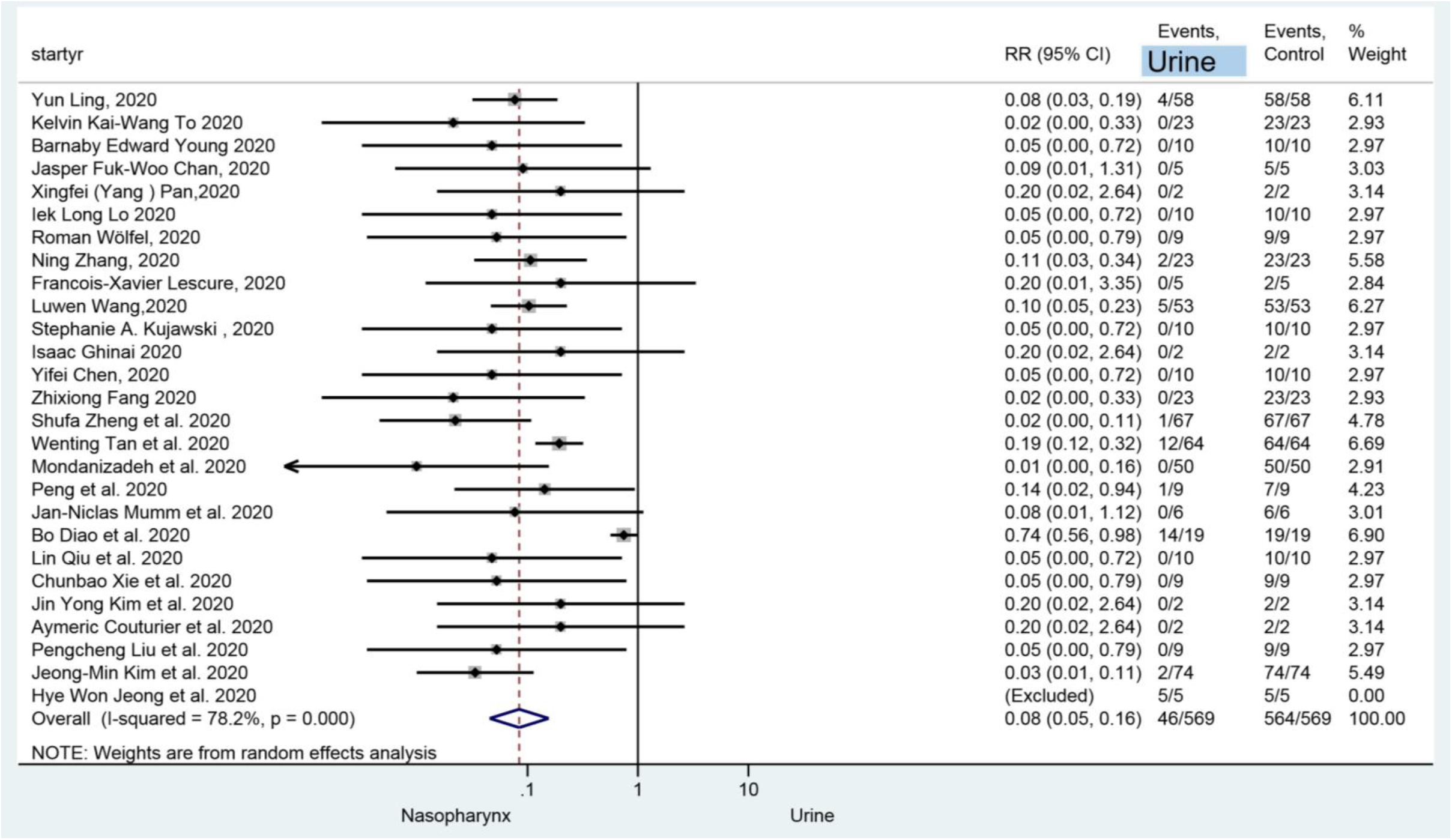
Forest plot, Relative risk of urine shedding of SARS-CoV-2 compared to pharyngeal specimens in the confirmed COVID-19 patients

**Figure 4:**
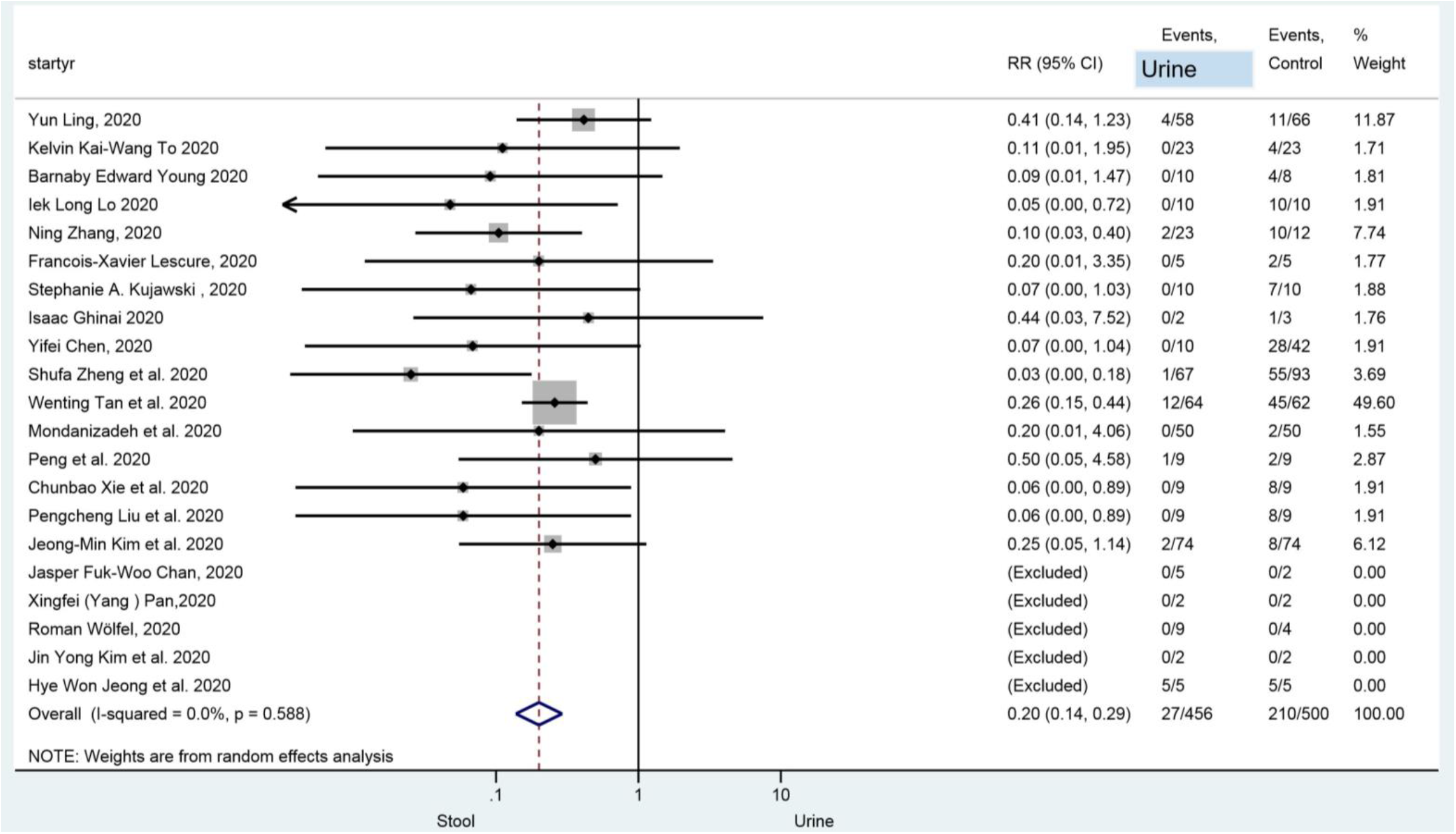
Forest plot, Relative risk (RR) of urine shedding of SARS-CoV-2 compared to the stool in the confirmed COVID-19 patients

**Figure 5:**
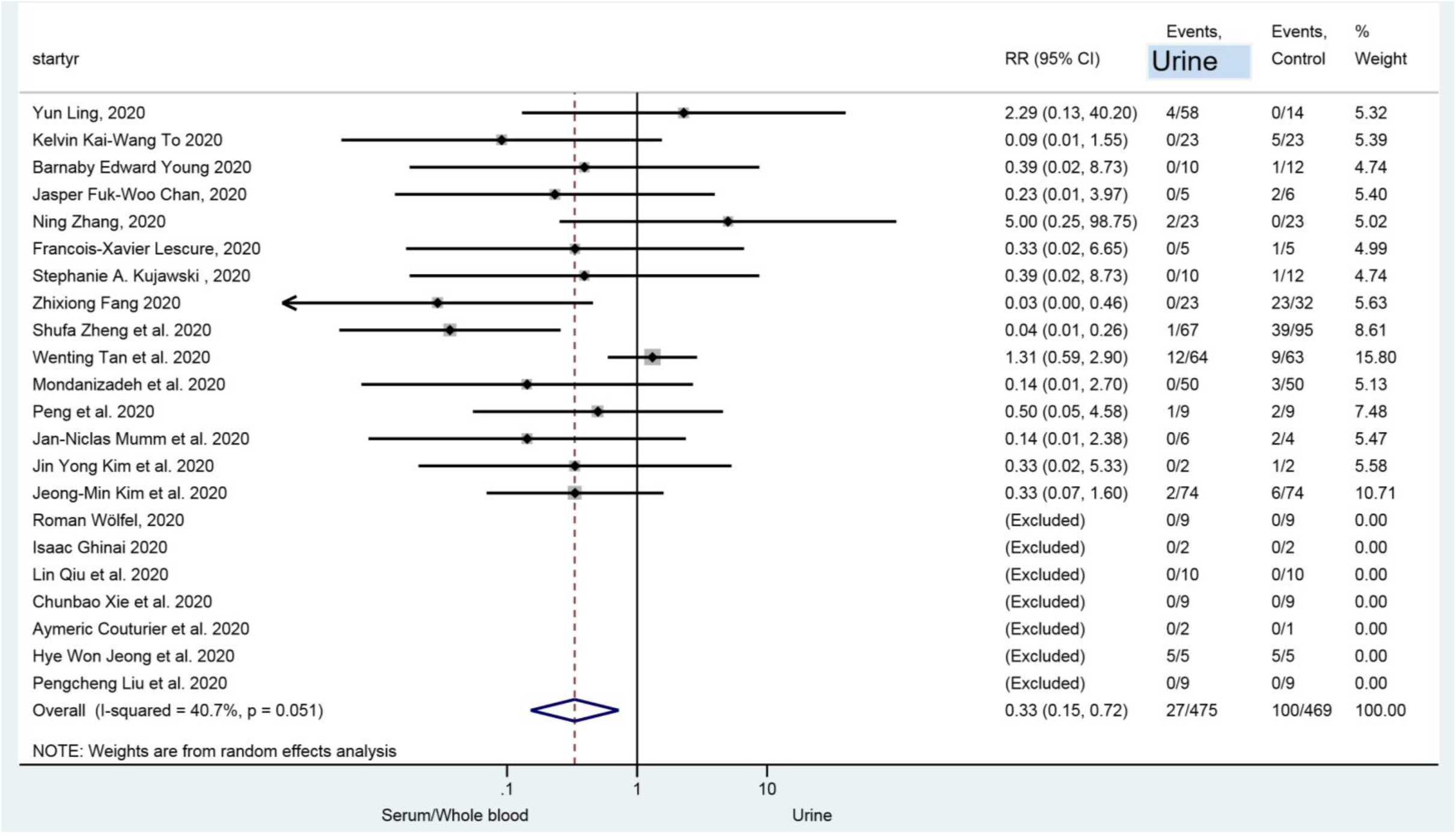
Forest plot, Relative risk of urine shedding of SARS-CoV-2 compared to blood in the confirmed COVID-19 patients

## Discussion

According to our three meta-analyses, stool and blood tests are associated with a significantly higher positive rate than urine (Figures 1-3). These results indicated that when the naso-/oro-pharyngeal SARS-CoV-2 test is negative, stool, and/or blood tests are more helpful for virus diagnosis than urine. Nevertheless, different factors such as the nature of viruses, missing data, design flaws, or methodological limitations might have contributed to these findings. The importance of urinary viral infection should not be ignored in terms of protective measures. We reviewed some important factors that can influence the results.

### Association Between Severity, Fatality, ICU admission rate, and Sepsis, with Urinary Positivity

COVID severity can be evidenced by the needs for mechanical ventilation, ICU care, as well as higher fatality. Viremia and sepsis are usually representative of more severe forms of diseases and a potential source for viral urinary shedding.

In SARS-CoV-1, viremia was reported to be associated with disease severity (61). Similarly, severely ill patients in the setting of SARS-CoV-2 disease may have augmented and prolonged the existence of the virus in blood and other body fluids (10). Furthermore, urinary viral positivity was found in severe cases in a study (7). SARS-CoV-2 viremia also may occur in patients with underlying comorbidities (62). Moreover, the duration of viral RNA excretion in respiratory and stool specimens may be longer in the cases treated with glucocorticoid, an immunosuppressive medication (8, 10). Accordingly, we emphasize the importance of inclusion of severely ill patients and ones with an underlying disease in the shedding studies.

In a study with a comparatively higher rate of urinary shedding (9.5%), a significantly higher proportion of the population had severe disease (40%) along with a case fatality of 6% (9). In another study, the duration of the existence of the virus in the blood of ICU patients was longer. The authors suggested its relationship with blood viral load and disease severity. Viruses were not found in the urine samples. Although their classification based on ICU admission was informative, the study didn’t have any information on whether the cases who ended in death -which might bear a higher urinary positive rate-hadn’t been excluded from the study (63). Similarly, some other studies with totally negative urinary results were limited to the populations with mild or moderate severity with null fatality (64, 65).

According to figure 2, the incidence risk of viral shedding in stool or blood was similar for both groups of studies with and without positive results. While trying to justify why a study has positive urinary results but another study doesn’t, the mentioned finding might not be in favor of the hypothesis that it could have arisen from systematic flaws in or in inclusion or exclusion criteria, sampling, or handling methods.

Nevertheless, a lack of concordance between the severity of disease and ICU admission rates in two groups in table 2 indicates heterogeneity of study populations among the studies.

As a result, since the difference in the severity of COVID-19 may contribute to the urinary negative results, we suggest to consider it while planning the study.

### Testing Frequency, Phases of the Disease, and Association with Urinary Results

Determining when the virus is detectable in the urine, is not simple. Different phases of the disease may lead to considerable differences in viral loads and peak concentrations and can be another factor responsible for different urinary findings. Toward the end of the period of the COVID disease, the virus is shown to be only intermittently detectable in pharyngeal swabs (41). Since the pharyngeal samples have much higher positive rates, finding a virus in the other fluid types could be more challenging.

As reported in SARS-CoV-1 studies, the urinary positivity rate was up to 42% at the end of the second week (5) with a peak occurring at weeks 3–4 and even shedding in the convalescent phase (66, 67). Similarly, for SARS-CoV-2, positive urine samples were detected at the latest available detection point (16 or 21 days after illness onset) (7).

Another study with a urinary positivity rate of 6.9% for viral RNA, revealed urine could stay positive after the throat swabs turned negative (8).

Although regular serial sampling was performed for pharyngeal specimens in some of the studies in our review, that was not the case for urine. Since repeat urinary testing is warranted especially in clinically suspected cases with an initially negative urinary result, we would emphasize the importance of systematic serial sample monitoring, throughout the disease phases, with an increased number of tested samples.

### Improper Exclusions, underpowered Sample size, and Bias in Urinary Results

Failure to find urinary viruses in many studies may be explained by their undersized study population. This may contribute to the positivity of urinary results found in studies done in China which were larger in population. Our review demonstrated that a considerable number of studies with a larger population were able to find positive urinary results (10, 34, 36, 53, 60, 68).

Moreover, there was a considerable difference in number between the population initially enrolled and finally tested for urine. Exclusions were not explained, as in some studies it was found to be more than half of the initial population. Except for one study, all the mentioned studies were accompanied by no positive urinary results (9, 49, 53, 69).

### Sample Handling and Laboratory Adjustments

Some studies with negative urinary results in this review had no stool positive findings (62, 65, 70), even though stool has been proven to have a high possibility of viral shedding (48.1%) (71). This co-negativity may also be explained by the errors in handling and laboratory technics.

Although real-time RT-PCR is considered as the gold standard for the diagnosis of COVID-19 (72), some factors such as the RNA quality, operator variability, or processing methods can affect the test results (73-75). Also, this technique does not distinguish between RNA residues and viable active viruses (76). As SARS-CoV-2 shares a high nucleotide identity with SARS-CoV-1 (82%) (77), using nonspecific real-time RT-PCR (e.g. SYBR Green method) may cause false-positive results. Some studies reviewed herein, lack information concerning the real-time RT-PCR type, primer, and probe sequences, candidate genes for virus detection, presence or absence of positive control, and the cycling parameters for PCR assay.

Real-time RT-PCR Ct (cycle threshold) values may also differ because of specimen collection or handling. The presence of several enzymes such as protease, RNase, or bacteria and the absence of proteins that stabilize RNA and virus in the urine may explain the quick degradation of viral RNA (78, 79). Technical improvement in the sampling to prevent degradation of the urinary viral RNA (such as immediate addition of lysis buffer to the fresh urine) may help to increase the diagnostic sensitivity and diminish false negative (78, 80). Further studies are needed to assess the efficacy of these methods in SARS-CoV-2.

Another source of false-positive urinary results can be passive contamination of urine samples with stool or other sources that can occur in severely ill or in the presence of diarrhea. The presence of different genotypes in urine or the comparatively higher RNA concentrations in urine would indicate active replication in the urine rather than contamination and spillage.

As can be noticed so far, this review encountered several limitations that resulted from a lack of high-quality evidence. We just mentioned the most important topics that help in building researches with a deeper focus on the design and methodologic quality in the future and help assess the viral shedding in urine and other specimens more efficiently. (Take-home bullet points)

## Conclusion

Based on review of the present literature, shedding of SARS-CoV-2 occurs in around 10 percent of population and it may have an association with the severity of systemic disease, need to admission in ICU and fatality. Investigating relationship between urine results and other factors is hardly possible without avoiding inappropriate exclusions. Furthermore, our review suggests that a larger population size may reveal more positive urinary cases. Moreover, using standardized laboratory quantitative control in real-time RT-PCR as well as repeat urinary testing would be warranted especially in patients with initially negative urinary results. (Take-home bullet points)

## Data Availability

This is a systematic review and meta-analysis. All data and analysis have been included into the paper.

